# The impact of lockdown during the COVID-19 pandemic on mental and social health of children and adolescents

**DOI:** 10.1101/2020.11.02.20224667

**Authors:** Michiel A. J. Luijten, Maud M. van Muilekom, Lorynn Teela, Hedy A. van Oers, Caroline B. Terwee, Josjan Zijlmans, Leonie Klaufus, Arne Popma, Kim J. Oostrom, Tinca J. C. Polderman, Lotte Haverman

## Abstract

**Importance:** It is unknown how a lockdown during the COVID-19 pandemic impacts children’s and adolescents’ mental and social health.

**Objective:** To compare mental and social health of children and adolescents during the COVID-19 lockdown versus before, identify associated factors, describe the change in atmosphere at home and qualitatively assess the impact of COVID-19 regulations on daily life.

**Design:** Cross-sectional study comparing two Dutch representative samples of children and adolescents (8-18 years); before COVID-19 (Dec2017-July2018) and during the COVID-19 lockdown (April/May 2020).

**Setting:** Population-based

**Participants:** Children and adolescents aged 8-18 years (M=13.4, 47.4% male), representative of the Dutch population on key demographics.

**Exposure(s):** COVID-19 pandemic lockdown

**Main Outcome(s) and Measure(s):** Patient-Reported Outcomes Measurement Information System (PROMIS^®^) domains: Global Health, Peer Relationships, Anxiety, Depressive Symptoms, Anger and Sleep-Related Impairment. Single item on atmosphere at home and open question regarding the impact of the regulations on the child/adolescent’s daily life

**Results:** Children and adolescents reported significantly worse PROMIS T-scores on all domains (absolute mean difference range, 2.1-7.1; absolute 95% CI range, 1.3-7.9) during the COVID-19 lockdown as compared to before COVID-19. More children reported severe Anxiety (during 16.7% vs. before 8.6%; relative risk 1.95; 95% CI 1.55-2.46) and Sleep-Related Impairment (during 11.5% vs. before 6.1%; relative risk 1.89; 95% CI 1.29-2.78). Fewer children reported poor Global Health (during 1.7 vs. before 4.6%; relative risk 0.36; 95% CI 0.20-0.65). More mental and social health complaints during the COVID-19 lockdown were found in children and adolescents growing up in a single-parent family, having ≥three children in the family, a negative change in work situation of parents due to COVID-19 regulations, and having a relative/friend infected with COVID-19. A small effect was found on atmosphere at home during the lockdown compared to before (mean difference, −3.1; 95% CI, −4.1 −−2.1). A large majority (>90%) reported a negative impact of the COVID-19 regulations on their daily life.

**Conclusions and Relevance:** This study showed that governmental regulations regarding lockdown pose a serious mental and social health threat on children and adolescents that should be brought to the forefront of political decision making and mental health care policy, intervention and prevention.

**Key points:** *Question:* What is the impact of lockdown during the COVID-19 pandemic on mental and social health in children and adolescents compared to before COVID-19?

*Findings:* This population-based study shows that during the COVID-19 lockdown children and adolescents report lower mental and social health, especially on anxiety and depressive symptoms.

*Meaning:* In proposing new COVID regulations (e.g., closing schools) governments should be mindful of the negative impact of a lockdown on mental and social health of children and adolescents.

## Introduction

The COVID-19 pandemic has an enormous impact on society as a whole, and certainly also on children and adolescents. Although children and adolescents are less affected by morbidity and mortality^1^, the restrictions imposed by governments worldwide profoundly impact their daily life, including their mental and social health.^2^

In the Netherlands, the first COVID-19 patient was identified on February 27^th^ 2020 and restrictions were imposed by the government starting on March 12^th^ 2020. People were asked to stay inside and work from home as much as possible, to comply to social distancing (1.5 meter) and all large events were canceled. On March 15^th^, a ‘partial’ lockdown was implemented.^3^ All schools and child care facilities were closed (unless one or both parents had an occupation classified as essential), as well as sports and leisure facilities, bars, and restaurants. However, children were still allowed to play outside, and visitors up to three persons at home were permitted. On May 11^th^ primary schools were partially reopened and on June 2^nd^ secondary schools followed.

During the lockdown, children and adolescents were experiencing physical isolation from their classmates, friends, teachers, and other important adults (e.g., grandparents). This might not only result in feelings of loneliness, but could potentially lead to precarious situations for children from unsafe domestic situations, due to a lack of escape possibilities. In addition, children and adolescents may experience mental health problems due to the COVID-19 pandemic itself, such as increased anxiety, as they might fear that they or their loved ones will get infected or they might worry about the future of the world.

Several cross-sectional and longitudinal studies on the effects of the COVID-19 pandemic on mental health in adults have now been published. Increased levels of anxiety, depression, suicidal ideation and (post-traumatic) stress, decreased psychological well-being, and a high percentage of sleep problems have been reported.^4-13^ Poor mental health was associated with female gender, younger age, low educational level, living alone/being divorced, having no work, low income, a socially disadvantaged background, and having an infected relative with COVID-19.^4-12^

However, studies on the effects of the COVID-19 lockdown on social and mental health of children and adolescents are yet sparse. Several opinion papers prelude on the most probable effects, and they have painted an image of loneliness, anxiety and depression following social isolation.^14,15^ Additionally, these papers have described increased tension at home and child abuse as possible consequences during the lockdown.^16,17^ Several authors fear that the lockdown will magnify existing health disparities and that certain communities (e.g., with migrant background and low socioeconomic status) will be more vulnerable to develop mental and social health issues.^15,18,19^ Notably, it has been hypothesized that some children may also experience positive consequences of the lockdown such as personal and social alleviation, family cohesion and reduction of stress.^20^

One systematic review on the effects of previous pandemics on mental health of children and adolescents is available, indicating that social isolation and quarantining has a negative impact on anxiety, depressive and fear symptoms.^2^ Additionally, only three cross-sectional survey studies are currently available, all from China, focusing on the impact of the COVID-19 pandemic and lockdown specifically. They reported prevalences of anxiety and depressive symptoms of 19% and 23% for primary school children,^21^ of 37% and 44% for high school children,^22^ and clinically elevated depressive symptoms scores for >22% of children and adolescents^23^ during the COVID-19 lockdown. These prevalences were reported to be higher compared to pre-COVID-19 established cut-offs and percentages in China, however none of these differences were statistically tested. Especially girls, older children, children living in an urban region and children having a COVID-19 infected friend/relative appeared to be most prone to mental health problems.^22,23^

A better understanding of how the governmental restrictions during the COVID-19 pandemic affect children’s and adolescents’ mental and social health can help guide future interventions and inform policy makers. The current study compares mental and social health of a representative sample of Dutch children and adolescents during the COVID-19 lockdown to earlier collected before COVID-19 reference data. The aims of this study were to: (1) quantify differences in mental and social health of children and adolescents before and during the COVID-19 lockdown, (2) identify factors that are associated with poorer mental and social health during the COVID-19 lockdown, (3) examine the change in overall atmosphere at home before and during the COVID-19 lockdown, and (4) qualitatively assess the impact of the COVID-19 lockdown on their daily life of children and adolescents.

## Methods

### Participants and procedure

#### Before COVID-19

As part of a larger Patient-Reported Outcome Measurement Information System (PROMIS) validation study, two studies were conducted between December 2017 and July 2018 in the Dutch general population to collect representative data of children and adolescents (8-18 years) on physical, mental and social aspects of health. A two-step random stratified sampling method was used to ensure representativeness on key demographics. Parents participating in existing panels were approached by two independent online research agencies (‘Kantar Public’ or ‘Panel Inzicht’). Both panels consist of families living across the Netherlands, that provided informed consent to be approached through e-mail for completing questionnaires for a small financial compensation. Children were subsequently approached by their parents to complete self-report questionnaires. Children completed the questionnaires through our research website of the KLIK Patient-Reported Outcome Measures (PROM) portal^24^ or the panel website. Parents were asked to complete a socio-demographic questionnaire. All children and parents provided informed consent and the studies were approved by the Medical Ethics Committee Amsterdam UMC. The samples were representative of the Dutch general population within 2.5% on most key demographics (age, gender, ethnicity, region, and educational level) compared to population numbers in 2017. Parental country of birth and parental educational level data were differently categorized for PROMIS Anxiety and Depressive Symptoms and therefore not useable for this study.^25^

#### During COVID-19

During the COVID-19 lockdown, between April 10^th^ and May 5^th^ 2020, data were collected by ‘Panel Inzicht’ from another sample of children and adolescents from the Dutch general population. The aim was to collect data on the same measures in a representative sample of approximately 1000 children with similar characteristics (within 2.5% of the previously mentioned key demographics) as the before COVID-19 sample. Data collection procedures were similar as in 2018, with the addition of a few COVID-19 specific questions for children, adolescents and parents. Children and parents provided informed consent and the study was approved by the Medical Ethics Committee Amsterdam UMC.

### Measures

#### Socio demographic questionnaires

Parents completed questions about themselves (region of residence, country of birth, educational level, marital status, and number of children) and their child (age, gender).

#### PROMIS pediatric measures

PROMIS item banks and scales were developed and validated, to measure generic unidimensional domains (e.g., anxiety) of physical, social or mental health using modern psychometric techniques.^26^ The item banks can be administered as Computerized Adaptive Test (CAT), where items are selected based on responses to previously completed items, resulting in a reliable score with a few items.^27^ Six Dutch-Flemish PROMIS pediatric measures (Scale V2.0 - Anger^28^, CAT V2.0 - Peer Relationships^29^, Scale V1.0 - Global health (7+2)^30^, CAT V1.0 - Sleep-related Impairment^31^, CAT V2.0 - Anxiety^32^, and CAT V2.0 - Depressive Symptoms^32^) were completed by children and adolescents. All PROMIS measures use a 7-day recall period, and items are scored on a five-point Likert scale. All items range from ‘never’ to ‘(almost) always’ except for Global Health, where response categories differ for each item (e.g. ‘excellent’ to ‘poor’). Total scores are calculated by transforming the item scores into a *T-score* which has a mean of 50 and standard deviation (SD) of 10 in the U.S. general population. For all measures higher scores represent more of the construct. For Anger (9 items) and Global Health (7+2 items) all items were administered. The US item parameters were used in the CAT algorithm and T-score calculations, as by PROMIS convention.

#### COVID-19–related additional questions

Three closed-ended questions were added for parents about whether there was a negative change in work situation of one of the parents/caregivers due to COVID-19 regulations (e.g., loss of income, reduced number of working hours, unemployment), whether a friend or relative had been infected with COVID-19 and if the child still attended child care/school during lockdown (e.g., both parents performing essential occupations).

Children and adolescents were asked to complete three COVID-19-specific questions: ‘How did you experience the atmosphere at home before the schools were closed?’ and ‘How do you experience the atmosphere at home now?’ rated on a visual analogue scale (VAS) ranging from 0 ‘Not pleasant at all’ to 100 ‘Very pleasant’, and an open-ended question ‘How are the corona-regulations for you?’.

### Statistical analyses

For all statistical analyses the Statistical Package for Social Sciences (SPSS), version 26.0, was used.

First, descriptive analyses (mean and percentages) were used to characterize the participants in the different samples. To compare samples, independent T-tests (for continuous variables) or chi-square tests of independence (for categorical variables) were performed.

Second, to test whether mental and social health of the sample during COVID-19 differed from the sample before COVID-19 on PROMIS T-scores, a one-way analysis of covariance (ANCOVA) was performed per PROMIS domain, adjusted for differences in sociodemographic characteristics. Mean differences (95% CI) were reported. A mean difference in domain T-scores of 0.25 SDs was considered meaningful.^33^

Percentages of children and adolescents reporting ‘severe’ symptoms or ‘poor’ functioning on the PROMIS measures before and during the COVID lockdown were compared. Severe symptoms or poor functioning was defined as a T-score 1.5 SD above or below the mean T-score before COVID-19 respectively, except for Peer Relationships, where 2 SD was used as cut-off for poor functioning (see www.HealthMeasures.net). Differences in proportions of severe scores were tested using chi-squares tests of independence and the relative risk (RR) with 95% confidence intervals (95% CI) were reported. The RR represents the risk of a child measured during the COVID-19 lockdown having a severe score compared to before COVID-19. A ratio >1 indicates more risk.

Third, to determine which variables were significantly associated with mental and social health during the COVID-19 lockdown, a multivariable linear regression analysis was performed for each PROMIS domain. The following variables were included as independent variables: age, gender, parental country of birth, marital status, region, number of children in the family, parental educational level, change in work situation due to COVID-19 regulations, infected relative/friend with COVID-19 and if the child still attended child care/school during lockdown. No multicollinearity was present between these variables (all correlations <.50). Per domain the effect size of all independent variables was reported, expressed as unstandardized regression coefficient B (95% CI).

Fourth, to investigate changes in atmosphere at home a paired T-test was used to test the difference between the two single items. Mean difference (95% CI) was reported.

Fifth, to assess the impact of the COVID-19 lockdown on the daily life of children, the open ended question ‘How are the corona-regulations for you?’ was qualitatively analyzed (by LT and HAvO) using thematic analysis.^34^ The answers were categorized into positive, neutral or negative experiences and thereafter clustered into themes. Themes were ranked according to their frequency of occurrence (high to low).

## Results

### Sociodemographic characteristics

During the COVID-19 lockdown, 844 children participated. This sample showed similar characteristics (most variables within 2.5% from each other) compared to the sample collected before COVID-19 (total N=2401). Significant differences were found on four variables (Table 1); age (during M=13.4 (SD=2.80) versus before M=13.1 (SD=3.14), mean difference=0.3; 95% CI, −0.54 – −0.06), both parents born in the Netherlands (during 88.2% versus before 79.8%, χ^2^(1)=29.884, p<0.001), parents with a low educational level (during 9.0% versus before 12.8%, χ^2^(2)=7.470, p=0.024), and families with one child (during 25.5% versus before 15.5%, χ^2^(2)=44.728, p<0.001).

**Table 1.**
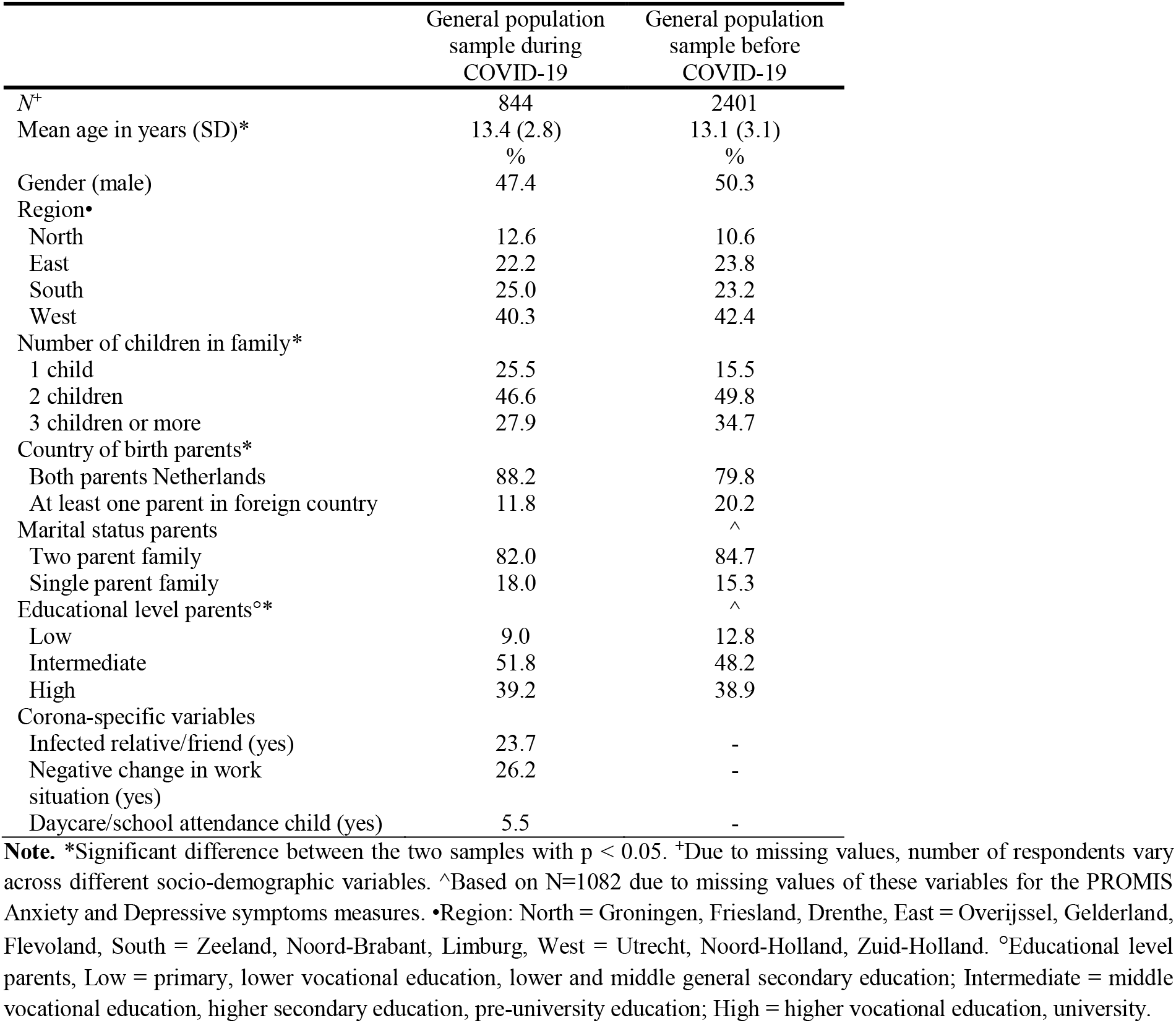
Socio-demographic characteristics of participants per group.

|  | General population<br>sample during<br>COVID-19 | General population<br>sample before<br>COVID-19 |
| --- | --- | --- |
| N<sup>+</sup> | 844 | 2401 |
| Mean age in years (SD)* | 13.4 (2.8) | 13.1 (3.1) |
|  | % | % |
| Gender (male) | 47.4 | 50.3 |
| Region<sup>•</sup> |  |  |
| North | 12.6 | 10.6 |
| East | 22.2 | 23.8 |
| South | 25.0 | 23.2 |
| West | 40.3 | 42.4 |
| Number of children in family* |  |  |
| 1 child | 25.5 | 15.5 |
| 2 children | 46.6 | 49.8 |
| 3 children or more | 27.9 | 34.7 |
| Country of birth parents* |  |  |
| Both parents Netherlands | 88.2 | 79.8 |
| At least one parent in foreign country | 11.8 | 20.2 |
| Marital status parents |  | <sup>^</sup> |
| Two parent family | 82.0 | 84.7 |
| Single parent family | 18.0 | 15.3 |
| Educational level parents<sup>°</sup>* |  | <sup>^</sup> |
| Low | 9.0 | 12.8 |
| Intermediate | 51.8 | 48.2 |
| High | 39.2 | 38.9 |
| Corona-specific variables |  |  |
| Infected relative/friend (yes) | 23.7 | - |
| Negative change in work situation (yes) | 26.2 | - |
| Daycare/school attendance child (yes) | 5.5 | - |

**Note.** \*Significant difference between the two samples with $p < 0.05$ . <sup>+</sup>Due to missing values, number of respondents vary across different socio-demographic variables. <sup>^</sup>Based on N=1082 due to missing values of these variables for the PROMIS Anxiety and Depressive symptoms measures. <sup>•</sup>Region: North = Groningen, Friesland, Drenthe, East = Overijssel, Gelderland, Flevoland, South = Zeeland, Noord-Brabant, Limburg, West = Utrecht, Noord-Holland, Zuid-Holland. <sup>°</sup>Educational level parents, Low = primary, lower vocational education, lower and middle general secondary education; Intermediate = middle vocational education, higher secondary education, pre-university education; High = higher vocational education, university.

### Differences in mental and social health in children and adolescents during versus before the COVID-19 lockdown

During the COVID-19 lockdown, children and adolescents reported worse T-scores than children and adolescents before the COVID-19 lockdown on all PROMIS domains, after controlling for age, parental country of birth, parental educational level and number of children (absolute mean difference range, 2.06–7.05; absolute 95% CI range, 1.25–7.86) (Table 2). Largest differences were found for Anxiety (mean difference=7.1, 95% CI, 6.2-7.9) and Depressive Symptoms (mean difference=4.9; 95% CI, 4.0– 5.7).

**Table 2.** Mean PROMIS T-scores and significant mean differences in the general population during and before COVID-19, adjusted for age, parental country of birth, parental educational level and number of children.

| | General population sample during COVID-19 | | | General population sample before COVID-19 | | | *p* * | $\eta^2$ | Mean difference (95% CI)^ |
| --- | --- | --- | --- | --- | --- | --- | --- | --- | --- |
|  | *N* | *M* | *SD* | *N* | *M* | *SD* |  |  |  |
| PROMIS Global Health<sup>+</sup> | 813 | 46.2 | 6.9 | 1082 | 48.3 | 9.8 | <0.01 | 0.01 | -2.1 (-2.9 – -1.3) |
| PROMIS Peer Relationships<sup>+</sup> | 813 | 44.3 | 7.0 | 527 | 46.9 | 9.5 | <0.01 | 0.02 | -2.6 (-3.5 – -1.7) |
| PROMIS Anxiety<sup>•</sup> | 813 | 50.5 | 7.6 | 1318 | 43.8 | 9.7 | <0.01 | 0.12 | 7.1 (6.2 – 7.9)<sup>°</sup> |
| PROMIS Depressive Symptoms<sup>•</sup> | 813 | 49.4 | 8.0 | 1318 | 44.7 | 10.6 | <0.01 | 0.05 | 4.9 (4.0 – 5.7) |
| PROMIS Anger<sup>•</sup> | 813 | 47.3 | 8.2 | 527 | 44.2 | 11.4 | <0.01 | 0.02 | 3.1 (2.0 – 4.1) |
| PROMIS Sleep Related Impairment<sup>•</sup> | 813 | 49.9 | 8.7 | 527 | 47.6 | 10.0 | <0.01 | 0.02 | 2.5 (1.5 – 3.5) |

**Note.** CI = confidence interval. <sup>+</sup>Higher scores indicate better functioning. <sup>•</sup>Higher scores indicate more symptoms. <sup>\*</sup>*P*-value of the main effect of the ANCOVA. <sup>°</sup>Not corrected for parental educational level due to missing values. <sup>^</sup>Adjusted mean differences and CI.

Significantly more children reported severe Anxiety (during 16.7% versus before 8.6%; RR, 1.95; 95% CI, 1.55–2.46) and severe Sleep-Related Impairment (during 11.5% versus before 6.1%; RR, 1.89; 95% CI 1.29–2.78) during the COVID-19 lockdown than before COVID-19 (Table 3). Fewer children reported poor Global Health (during 1.7% versus before 4.6%; RR, 0.36; 95% CI, 0.20–0.65).

**Table 3.** Percentage of participants with poor functioning or severe symptoms (>1.5 SD)° on the PROMIS domains for both samples *.

|  | *<br>General population sample |  | Relative risk (95% CI) |
| --- | --- | --- | --- |
|  | during COVID-19 | before COVID-19 |  |
|  | % | % |  |
| PROMIS Global Health* | 1.7 | 4.6 | 0.36 (0.20 – 0.65) |
| PROMIS Peer Relationships | 1.9 | 1.9 | 0.99 (0.46 – 2.19) |
| PROMIS Anxiety* | 16.7 | 8.6 | 1.95 (1.55 – 2.45) |
| PROMIS Depressive Symptoms | 7.1 | 8.2 | 0.87 (0.64 – 1.18) |
| PROMIS Anger | 3.7 | 5.5 | 0.67 (0.41 – 1.09) |
| PROMIS Sleep Related Impairment* | 11.5 | 6.1 | 1.89 (1.29 – 2.78) |

**Note.** CI = Confidence interval. \*p-value < 0.01. °For Peer Relationships a cut-off of 2 SD was used, as per HealthMeasures guidelines.

### Variables associated with poor mental and social health in children and adolescents during the COVID-19 lockdown

Lower Global Health was associated with a single-parent family (B=-3.00; 95% CI, −4.23 – −1.76). Lower Peer Relationships were reported by boys compared to girls (B=-1.25; 95% CI, −2.23 – −0.27). Increased Anxiety was associated with age (B=-0.34; 95% CI, −0.53 – −0.15), a single-parent family (B=1.46; 95% CI, 0.11–2.81), an infected relative or friend (B=1.94; 95% CI, 0.72–3.16) and parents with a negative change in work (B=3.01; 95% CI, 1.84–4.18). More Depressive Symptoms was associated highly educated parents (where intermediate differed from lower; B=2.24; 95% CI, 0.23– 4.24) and parents with a negative change in work situation (B=2.45; 95% CI, 1.20–3.70). More Anger was associated with age (B=-0.47; 95% CI, −0.67 – −0.28), highly educated parents (where intermediate differed from lower; B=2.30, 95% CI, 0.27–4.33 and high differed from lower; B=2.10; 95% CI, 0.01– 4.19), three or more children (B=2.07; 95% CI, 0.52–3.62) and parents with a negative change in work situation (B=1.71; 95% CI, 0.45–2.98). Finally, more Sleep-Related Impairment was related to the country of birth of parents (≥ one foreign parent; B=1.95; 95% CI, 0.13–3.77), a single parent family (B=2.07; 95% CI, 0.53–3.62) and parents with a negative change in work (B=2.53; 95% CI, 1.19–3.87) (Table 4).

**Table 4.** Variables associated with mental and social health in children and adolescents during the COVID-19 lockdown.

|  | Global Health<sup>+</sup> | Peer Relationships<sup>+</sup> | Anxiety<sup>•</sup> | Depressive Symptoms<sup>•</sup> | Anger<sup>•</sup> | Sleep-Related Impairment<sup>•</sup> |
| --- | --- | --- | --- | --- | --- | --- |
| Covariates | *B* | *B* | *B* | *B* | *B* | *B* |
| Age | -0.03 | 0.11 | -0.34** | -0.19 | -0.47** | -0.09 |
| Male | 0.50 | -1.25* | -0.32 | -0.56 | 0.12 | -1.11 |
| Region <sup>^</sup> |  |  |  |  |  |  |
| West | Ref | Ref | Ref | Ref | Ref | Ref |
| North | -0.22 | -0.19 | -0.50 | -1.18 | -0.97 | -0.52 |
| East | -0.88 | -0.07 | 0.32 | 0.21 | -0.17 | 0.38 |
| South | -0.78 | -0.32 | -0.03 | -0.69 | -0.74 | -0.94 |
| Foreign country of birth parents | -0.33 | -0.60 | 0.45 | 0.32 | 0.37 | 1.95* |
| Educational level parents<sup>°</sup> |  |  |  |  |  |  |
| Low | Ref | Ref | Ref | Ref | Ref | Ref |
| Intermediate | -0.68 | 0.78 | 1.60 | 2.24* | 2.30* | 1.19 |
| High | 0.87 | 1.51 | 1.12 | 1.94 | 2.10* | 0.02 |
| Single parent family | -3.00** | -0.76 | 1.46* | 1.02 | 0.91 | 2.07** |
| Number of children in family |  |  |  |  |  |  |
| 1 child | Ref | Ref | Ref | Ref | Ref | Ref |
| 2 children | 0.37 | -0.81 | 0.31 | 0.08 | 1.29 | 0.11 |
| 3 or more children | 0.60 | 0.16 | -0.14 | -0.17 | 2.07** | 1.26 |
| Negative change in work situation | -0.88 | -0.46 | 3.01** | 2.45** | 1.71** | 2.53** |
| Infected relative/friend with COVID-19 | 0.96 | 0.84 | 1.94** | 1.11 | 0.11 | 0.99 |
| Daycare/school attendance child | -0.65 | 0.20 | 1.20 | 2.04 | 1.15 | 2.73 |

**Note.** *B* = Unstandardized regression coefficient of multivariable linear regression model. \*\*p-value <.01, \*p-value <.05. <sup>+</sup>Higher scores indicate better functioning. <sup>•</sup>Higher scores indicate more symptoms. <sup>°</sup>Educational level parents divided into three categories: Low = primary, lower vocational education, lower and middle general secondary education; Intermediate = middle vocational education, higher secondary education, pre-university education; High = higher vocational education, university. <sup>^</sup>Region: North = Groningen, Friesland, Drenthe, East = Overijssel, Gelderland, Flevoland, South = Zeeland, Noord-Brabant, Limburg, West = Utrecht, Noord-Holland, Zuid-Holland.

### Changes in atmosphere at home during the COVID-19 lockdown

Children and adolescents reported a worse atmosphere (mean difference=-3.1; 95% CI, −4.1 – −2.1) at home during the COVID-19 lockdown (M=78.2, SD=17.9) than before COVID-19 (M=81.4, SD=16.0).

### Impact of COVID-19 regulations on the daily life of children

The majority (∼90%) of children indicated that the COVID-19 lockdown had a negative impact on their daily life. The most often mentioned issues (>50 children) were: 1) missing contact with friends, 2) not allowed to go to school, 3) missing freedom, 4) not allowed to participate in sports, 5) missing joyful activities (e.g., birthdays, holidays, parties, shopping), 6) difficulties with homeschooling 7) missing extended family, and 8) boredom (Table 5). A minority of children did not experience any difficulties with the COVID-19 lockdown regulations (∼7%) (e.g., *‘It does not bother me’*) or reported positive consequences (∼3%) (e.g., *‘I really like that I can play with children in my neighborhood all day long’)*.

**Table 5.**
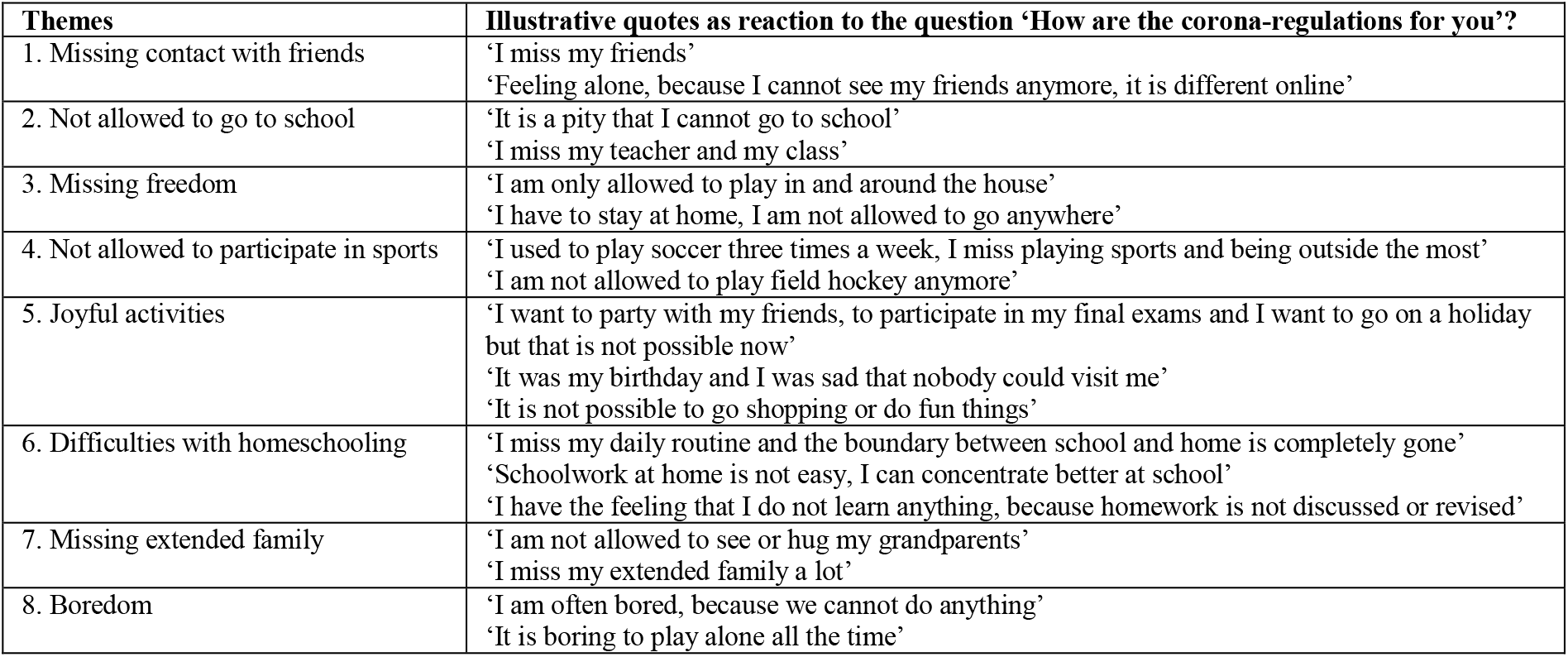
Themes regarding the negative impact of COVID-19 regulations ranked according to their frequency of occurrence (high to low).

## Discussion

This study compared the mental and social health of a representative sample of children and adolescents from the general population *during* the COVID-19 lockdown to a similar sample of children and adolescents *before* COVID-19. Children and adolescents reported poorer mental and social health during the COVID-19 lockdown on all six PROMIS domains. Substantial differences in percentages of children reporting severe anxiety and sleep-related impairments were observed. Fewer children and adolescents reported a poor global health during the COVID-19 lockdown, although the mean global health score was lower in this sample as compared to the sample before COVID-19. Significant associations with mental and social health complaints during the COVID-19 lockdown were found for family composition (growing up in a single-parent family or having three or more children in the family), a negative change in work situation of parents due to COVID-19 regulations, and an infected relative/friend with COVID-19. Children and adolescents reported a small decrease in atmosphere at home during the lockdown. The majority of children and adolescents revealed a negative impact of the COVID-19 regulations on their daily life, that far outweigh the number of children who reported a positive effect. Especially missing contact with friends was considered important.

The results of this study confirm the suspicions of child and youth care professionals that the COVID-19 lockdown has negative effects on mental and social health of children and adolescents. In opinion papers, professionals elaborated on the vulnerability of this group and expected more feelings of loneliness, anxiety and depression, as well as a more tense atmosphere at home.^14-17^ Concerns were also expressed that the COVID-19 lockdown would lead to an increase in inequality and that children and families with lower social economic status would be more susceptible to mental health issues.^15,18,19^ Although this study could not definitely confirm these concerns, children from single parent families, from families with three or more children, and with parents who had a negative change in work situation reported more mental and social health problems during the COVID-19 lockdown.

While the Dutch partial lockdown was substantially different from the Chinese full lockdown, our results are in line with the three studies from China,^21-23^ who also reported higher anxiety and depressive symptom during the COVID-19 lockdown. Likewise, one of the Chinese studies also found that having an infected relative/friend with COVID-19 was predictive of more anxiety.^23^ In addition to the effects on anxiety and depressive symptoms, our study results show negative effects of the COVID-19 lockdown on anger, sleep-related impairment and peer relationships.

Although the mean T-score on Global Health was lower (worse) in the sample during the lockdown as compared to before COVID-19, a lower percentage of children and adolescents reported poor Global Health during COVID-19. This finding is puzzling and prompts further study in a follow-up design that is underway in our department.

Some limitations of this study need to be taken into account. Although the aim was to obtain two representative samples that were completely comparable, small differences were found on age, parental country of birth and educational level, and family composition. However, these differences were corrected for in our comparisons between the groups. In addition, the data collection during COVID-19 took place in April and May (2020), whereas the study data collection before COVID on anxiety and depression mainly took place in January and February (2018). Worse mental health is often reported during winter times.^35^ This difference could have led to an underestimation of the actual impact of the COVID-19 lockdown.

We found that children and adolescents from families with certain risk factors (e.g. single parent families) are more vulnerable to mental and social health problems. These children and adolescents should be in sight of health care professionals. However, in this study children and adolescents with existing mental or somatic problems were not included, while it is conceivable that these groups are even more vulnerable. More research is needed to study the mental and social health of these groups as well as to gain insight in the longitudinal effects, and to clarify if lower mental and social health scores are mainly due to the COVID-19 pandemic or the lockdown.

During the finalization of this paper, the Netherlands, and many other countries, are facing a second COVID-19 wave. This study showed that governmental regulations regarding lockdown pose a serious mental and social health threat on children and adolescents that should be brought to the forefront of political decision making and mental health care policy, intervention and prevention.

## Supporting information

STROBE Checklist

## Data Availability

Data may be made available upon request. Please contact the corresponding author.

## Acknowledgement section

### Authors’ contributions

LH conceived the study. MAJL, MMvM, LT, HAvO, and LH conceptualized and designed the study. MAJL, MMvM, LT, HAvO, CBT and LH developed the sociodemographic questionnaire and MAJL, MMvM and LT carried out the data collection. MMvM, MAJL, LT and JZ performed the statistical analyses and HAvO and LT performed the qualitative analyses. CBT supervised the statistical analyses. The first draft of the manuscript was written by MMvM, MAJL and LH, and all authors commented on previous versions of the manuscript. LH, HAvO, TJCP, AP, KJO and CBT supervised the process. All authors critically revised the manuscript for intellectual content and approved the final version.

### Data availability

Data are available upon reasonable request.

### Funding

The data collection in this study was supported by Stichting Steun Emma Kinderziekenhuis, the Dutch National Health Care Institute and the Netherlands Organization for Health Research and Development.

### Ethics approval

The Medical Ethical Committee of the Amsterdam UMC (location AMC and VUMC) approved the protocol and judged that the Dutch Medical Research Involving Human Subjects Act does not apply to this study. All procedures performed were in accordance with the 1964 Helsinki declaration and its later amendments or comparable ethical standards.

### Previous presentation

Part of the data in this article has been presented as a poster and oral presentation during the annual International Society of Quality of Life Research (ISOQOL) that took place online from 20 to 23 of October 2020.

## Acknowledgements

We would like to acknowledge Stichting Steun Emma Kinderziekenhuis and the Dutch National Health Care Institute for their funding. Additionally, we would like to thank all children and adolescents that completed the questionnaires and Biomedia and Panel Inzicht for their support in setting up the research website and sending out the questionnaires.

